# Advanced Rock Climbers Exhibit Greater Finger Force and Resistance to Fatigue Compared to Novices During Treadwall Climbing

**DOI:** 10.1101/2020.04.27.20077560

**Authors:** Philip F. Ferrara, James Becker, John G. Seifert

## Abstract

**Background:** The purpose of the study was to investigate the effects of rock climbing experience on time to fatigue (TTF), finger flexor force production relative to body weight (REL FP), and changes in finger flexor muscle activity during tread wall climbing.

**Methods:** Eight advanced and seven novice sport rock climbers performed a climbing protocol on a treadwall system. The protocol consisted of climbing for six 5-minute intervals or until voluntary failure. A mounted force plate was used to measure finger force production before and after the climbing protocol. Subjects performed a 20-second maximum voluntary isometric contraction (MVIC) against the force plate with the dominant and non-dominant fingertips in a common climbing hand configuration known as a half crimp. Muscle activity was monitored with electromyographic electrodes placed bilaterally over the subject’s *flexor digitorum superficialis*. Median frequency (MF) and root mean square (RMS) were analyzed. The treadwall was set at a difficulty of 9 IRCRA/5.9 YDS. It rotated at 7 m/min and was fixed at 7° overhanging.

**Results:** Significant group differences were observed in subject characteristics, TTF (30±0 vs. 25.7±3.6 min), REL FP (5.6±1.2 vs. 3.2±0.9 N/kg BW), ΔFP (+1.5±12.3% vs. -31±16%), and ΔMF (+6.3±22.4% vs. -17.6±10.9%).

**Conclusion:** The results of this study demonstrate that advanced climbers possess greater finger flexor force production and resistance to fatigue compared to novices during bouts of climbing on a treadwall system. This may be attributed to physiological changes due to years of training, such as metabolic adaptations and oxygenation capacity in the forearm musculature.

## 1. Introduction

Rock climbing is a dynamic, multi-faceted sport with many specializations. One such variation, known as free climbing, involves using the hands, feet, and safety equipment to ascend a rock face.^1^ During a free climbing ascent, the finger flexors (such as the *flexor digitorum superficilias*, FDS), perform repeated isometric muscle contractions while the digits maintain tension against rock holds.^2^ These repetitive contractions slow blood flow to the forearm musculature, leading to ischemia, acidosis and localized muscular fatigue. Colloquially known as “forearm pump,” localized muscular fatigue is a primary cause of failure during a free climbing ascent.^3^ As climbing standards progressed, rock climbers began implementing specialized training tools to develop sport-specific strength and resistance to fatigue.^4^ These tools include the treadwall system, a motorized, rotating climbing wall with fixed plastic holds.

High finger force production relative to body weight (REL FP) is also an essential component of climbing, with an upward trend noted in trained participants.^4-7, 9-13^ Differences between trained climbers and novice/non-trained counterparts have been documented in the literature, including REL FP and finger flexor time to fatigue (TTF) during various tasks.^5-14^ While it does appear that trained climbers have greater REL FP and TTF, many of the previous studies used imitation climbing tasks to induce fatigue. This includes intermittent or sustained fingertip contractions against a force plate in a fixed body position. Currently, few studies have used actual climbing as a means of fatiguing subjects while monitoring acute changes in finger force production or muscle activity.^8^

Electromyography (EMG) is a means of measuring muscle activity via motor unit action potentials.^15^ Post-processing techniques such as median frequency (MF) and root meat square (RMS) have been used to verify shifts in motor unit composition and activation, respectively.^11, 15^ Previous climbing research implemented EMG to estimate changes in muscular activity of the finger flexors during fatiguing protocols.^11, 12, 16-18^ While previous literature suggests trained climbers have functional differences in fiber recruitment and a resistance to fatigue when compared to novices, these studies are again limited to isolated tasks as opposed to actual climbing.^12^

Current literature has established important physiological adaptations to years of high-level rock climbing, including changes in finger flexor force production, time to fatigue and muscle fiber recruitment patterns, however the protocols implemented oftentimes lack sport specificity. ^2, 3, 11, 12^ For example, Watts et al., 2008 found that the traditional method for measuring grip strength, the handgrip dynamometer, was not specific enough to measure muscle activation in common rock climbing hand configurations like the half crimp.^18^ Moreover, many previous protocols did not include any form of climbing to fatigue subjects. Thus, we have developed a new protocol to induce fatigue and assess changes in finger force production and muscle activation.

In the current study, a climbing-specific fatiguing protocol utilizes repeated bouts of climbing on a treadwall system to approximate climbing in the field. This protocol is used in combination with a climbing-specific finger strength testing device which has been adapted from the literature to measure change in finger flexor force production and muscle activation in a commonly-used hand configuration known as the half crimp.^5, 19^ This is the first time these tools have been used in conjunction, which allowed us to both assess fatigue patterns and measure change in force production in a climbing-specific manner.

The purpose of the present study is to investigate how rock climbing ability level affects time to fatigue, finger force production and muscle activation during intermittent bouts of tread wall climbing. It is hypothesized advanced rock climbers will have a greater time to fatigue, greater finger force production relative to body weight (REL FP), lesser change in finger force production (ΔFP), lesser change in median frequency (ΔMF) and lesser change in root mean square (ΔRMS) when compared to novice climbers.

## 2. Methods

### 2.1 Participant Selection

Subjects were selected for experience group (advanced vs. novice) based on self-described ability level, years of rock climbing experience, mean onsight and mean project grade as described by Draper et al.^20^ An “onsight” is defined as climbing a route within one attempt with no falls and no prior knowledge of the holds. A “project” is a route that is completed with no falls within five attempts but with prior knowledge of the holds.

Grading systems are used to measure the overall difficulty of a climbing route. The higher the grade, the more difficult the route and more skilled a climber needs to be to complete it without falling. All grades are reported in the International Rock Climbing Research Association’s universal scale (IRCRA), the American Yosemite Decimal System (YDS) and European French grading system where applicable. Both male and female advanced climbers were required to ^12^ For example, Watts et al., 2008 found that the traditional method for measuring grip strength, have at least five years of climbing experience. Advanced male subjects were required to climb a minimum 18 IRCRA/5.12a YDS/7a+ French.^21^ Advanced female subjects were required to climb a minimum rating of 15 IRCRA/5.11 YDS/6c French. Novice subjects were required to have at least one year of climbing experience and climb up to a rating of 10 IRCRA/5.10a YDS/5+ French. All subjects reported at least 50% of their training in the last six months be route climbing. This was done to ensure participants were conditioned to longer bouts of climbing. Prior approval of the study protocol was obtained from the Montana State University’s Institutional Review Board. All individuals provided informed consent prior to participating. Fifteen trained rock climbers participated in the study.

### 2.2 Experimental Design

This descriptive study used between-subject (advanced vs. novice) and within-subject (dominant vs. non-dominant hand) fixed measures with repeated measurements (pre-to post-exercise) to determine differences in finger flexor force production, endurance and muscle activity during tread wall climbing.

### 2.3 Experimental Procedure

#### 2.3.1 Instrumentation

To assess finger flexor force production in a climbing-specific hand configuration, a mounted force plate was adopted from Grant et al., 1996 (Fig. 1).^5^ The force plate (AMTI© MC3A-1000, Watertown, MA) was mounted to a reinforced wooden box with an adjustable elbow rest. All subjects were seated in 120°C of horizontal abduction at the shoulder and 90°C of elbow flexion, 0°C of wrist flexion, and 90°C flexion at the proximal interphalangeal joint.^5^ Joint angles were measured manually with a goniometer. The force plate was calibrated before each subject’s test using a fixed 5 gram weight. Prior to each MVIC, the force plate was tared to 0 ± 0.1 N. An electromyographic electrode (Delsys Trigno™, Delsys Inc., Nateck, MA, USA) was placed bilaterally over the belly of the FDS. To find the muscle belly, subjects were asked to flex the middle three digits at the proximal interphalangeal joint against an external force while the muscle was palpated, as per Blackwell et al.^22^ Hair was removed, the skin was abraded and cleansed with alcohol. Electrodes were placed over the muscle belly with respect to the pennation angle of the muscle.^15^ The electrodes were then fixed with sports tape and secured with athletic pre-wrap.

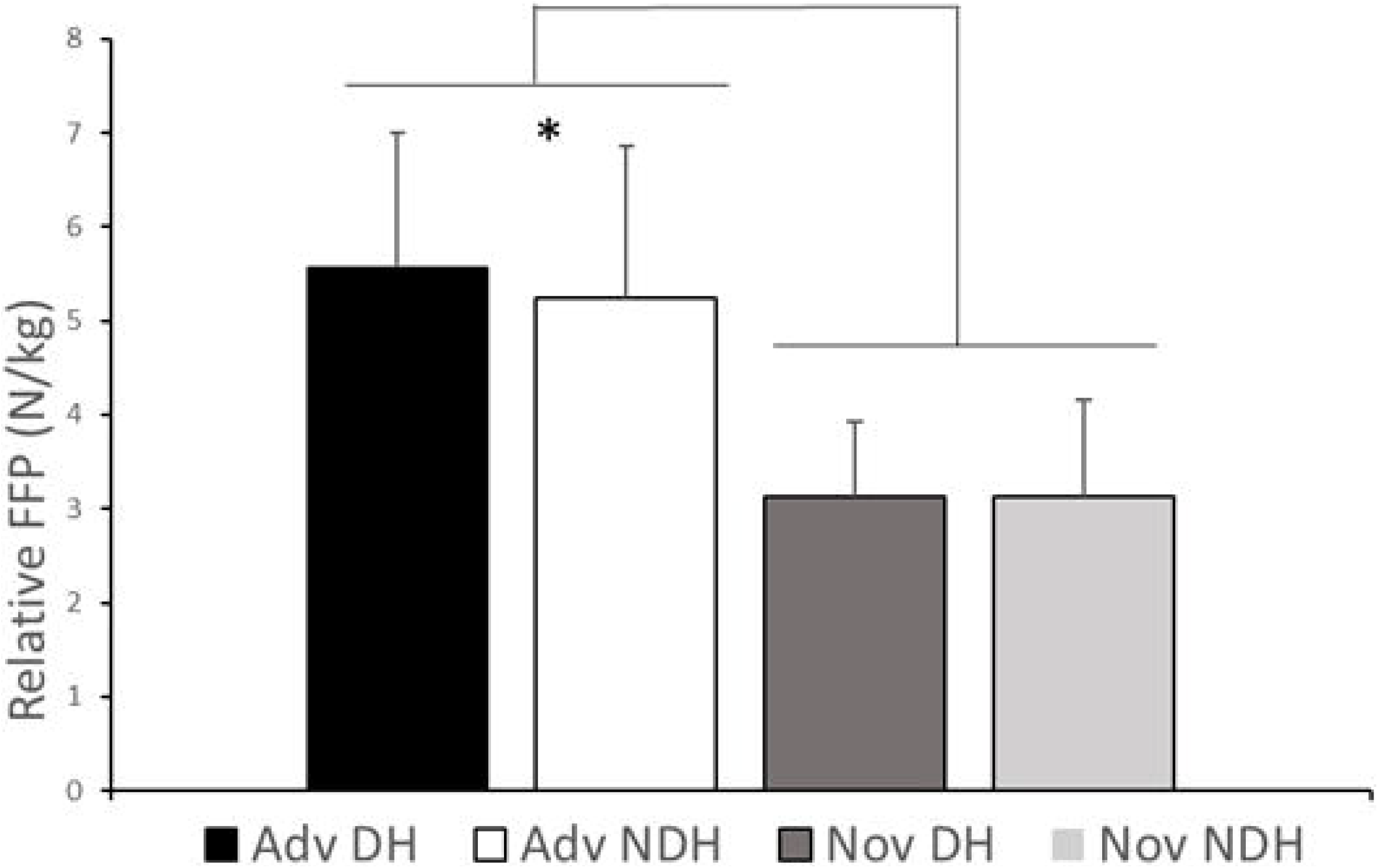

#### 2.3.2 Habituation and Warm-Up

A five-minute warm-up and habituation period were given to all participants, allowing for acclimation to the tread wall system (The Rock™, Ascent, Salt Lake City, UT, USA). Participants then completed a mock finger force test on the mounted force plate as a warm-up. The mock test involved two, 20-second submaximal isometric contractions against the force plate in the half crimp position with the dominant hand (DH) and non-dominant hand (NDH).^19^ A half crimp is a commonly-used hand configuration in climbing. The proximal interphalangeal joint is flexed to 90° while the finger tips apply force to rock holds. (see figure 1). During this mock test, joint angles and body positions were standardized across all subjects.

#### 2.3.3 MVIC Test Protocol

The MVIC test consisted of one 20-second maximal fingertip contraction in the half-crimp position with both the dominant and non-dominant hand. A countdown was given, at which point both force and EMG recording devices were triggered simultaneously. Verbal encouragement was given at the 10-, 15- and 18-second mark.

#### 2.3.4 Treadwall Protocol

All participants climbed the same treadwall route with no alternatives. The route was set with the assistance of USA Climbing-certified level 1 route setter at a rating of 9 IRCRA/5.9 YDS/5 French. Plastic holds were mirrored to fatigue both arms equally. The treadwall rotated at 7m/min and was set to 7°C overhanging to promote fatigue in the finger flexors.^23^ Subjects were allowed to drop one arm between holds, however they were not allowed to “shake out” during the test. A shake out is when the arms are alternately lifted over the head and shook to improve blood flow to the forearms.

Following the pre-exercise MVIC (MVIC_PRE_), subjects began the treadwall protocol. Subjects climbed for five-minute intervals, which mimicked the time a climber may spend on a route. Following the climbing interval, subjects immediately performed another MVIC test beginning with the dominant hand. No rest was given between DH and NDH. Finger strength tests were kept to less than two minutes to minimize recovery. Subjects then immediately climbed for another five minutes. This was repeated for a maximum of six sets or until subjects could no longer hang on to the holds (failure). Immediately after completion of the climbing protocol or at the point of volitional failure, the post-exercise MVIC was performed.

### 2.4 Data Analysis

Comparisons were made between the first (MVIC_PRE_) and fifth MVIC (MVIC_POST_) to account for differences in climb time. Force and EMG data were cropped to remove zeroes before and after the contraction. The remaining 20-second data set was cropped again, removing the first and last 10% (2 seconds) to account for transient phenomena.

#### 2.4.1 Force Data Processing

Force data was recorded with AMTI NetForce™ software and collected at 4096 Hz. Force data was analyzed using a custom written Matlab code (Mathwoks, Natick, MA, USA). Raw data was filtered with a zero-phase, low pass 2^nd^ order Butterworth filter with an 8 Hz cut-off.

The pre-exercise MVIC was used to determine each subject’s maximal force production. It was calculated as the mean of the filtered 16-second data set. Mean force production for each subsequent MVIC was divided by MVIC_PRE_ to normalize change in finger strength amongst all subjects over time.

#### 2.4.2 EMG Data Processing

Electromyographic data was recorded in Delsys EMGWorks® Acquisition software and collected at 1926 Hz. It was converted via a 16-bit A/D board (National Instruments USB-6225, Austin, Texas, USA). Electromyographic data was analyzed in Delsys EMGWorks® Analysis and filtered using a zero-phase, low pass 4^th^ order Butterworth filter with 300 Hz cut-off.^11^ Root mean square and MF was calculated on the filtered, cropped data. The RMS time window was 125 ms with a window overlap of 62.5 ms.

#### 2.5 Statistical Analysis

Unpaired t-tests were used to analyze subject characteristics and TTF. A 2×2×2 (group by hand by time) mixed analysis of variance (ANOVA) was performed to analyze force production during MVIC_PRE_ and MVIC_POST_. A 2×2 (group by hand) mixed ANOVA was performed to analyze ΔFP, ΔMF, and ΔRMS from MVIC_PRE_ to MVIC_POST_. Significance level was set at *p*≤*0.05 (*SPSS 25.0; SPSS Inc., Chicago, IL, USA). *Post hoc* comparisons were made using a Bonferroni procedure. All statistics are reported as mean ± standard deviation (SD). Data was stored in Excel® (Microsoft Corp., Redmond, WA, USA) spreadsheets.

## 3. Results

### 3.1 Subject Characteristics

Age (*p=0.001*), years of climbing experience (*p=0.001*), mean project grade (*p=0.001*) and mean onsight grade (*p=0.002*) were significantly different between advanced and novice participants. Subject characteristics reported and summarized in Table 1. Two novice subjects were excluded from data analysis due to premature muscular failure at six and eight minutes, respectively.

**Table 1.** Subject selection summary table (n=15)

| Subject | N | Age | Body Mass | Height | Years Experience | Mean Onsight Grade | Mean Project Grade |
| --- | --- | --- | --- | --- | --- | --- | --- |
| *Nov* | 7 | 21 ± 2.3* | 67.6 ± 3.8 | 67.6 ± 3.8 | 3.0 ± 2.6* | 10.0 ± 2.0* | 11.0 ± 1.0* |
| *Adv* | 8 | 29.3 ± 4.7 | 69.1 ± 6.9 | 69.1 ± 6.9 | 11.1 ± 5.2 | 20.0 ± 1.0 | 26.0 ± 1.0 |

\* = significant differences between groups at $p < 0.05$ . Reported as mean ± SD.
Climbing grades reported in the IRCRA rating scale.
Reported as mean ± SD.

### 3.2 Time to Fatigue

Time to fatigue was significantly longer for advanced climbers (30 ± 0 min) compared to the novice climbers (25.7 ± 3.6 min; *p=0.001*). All advanced subjects completed the climbing protocol. Only one novice subject completed the full, 30-minute protocol.

### 3.3 Relative Force Production

A significant group by hand interaction was observed for REL FP (*F*_1,1_*= 8.23, p*=*0.008*). *Post-hoc* analysis revealed the advanced groups’ DH (5.6 ± 1.2 N/kg BW) produced significantly more force than the advanced NDH (5.2 ± 1.3 N/kg BW, *F*_1_*= 11.78, p=0.004*). No difference was observed in the novice group’s DH (3.2 ± 0.8 N/kg BW) and NDH (3.2 ± 1.0 N/kg BW). Moreover, the advanced group produced significantly more force on average (5.4 ± 1.3 N/kg) when compared to the novice group (3.2 ± 0.9 N/kg BW, *F*_1_*= 39.6, p=0.001*). (Table 2)

**Table 2.**
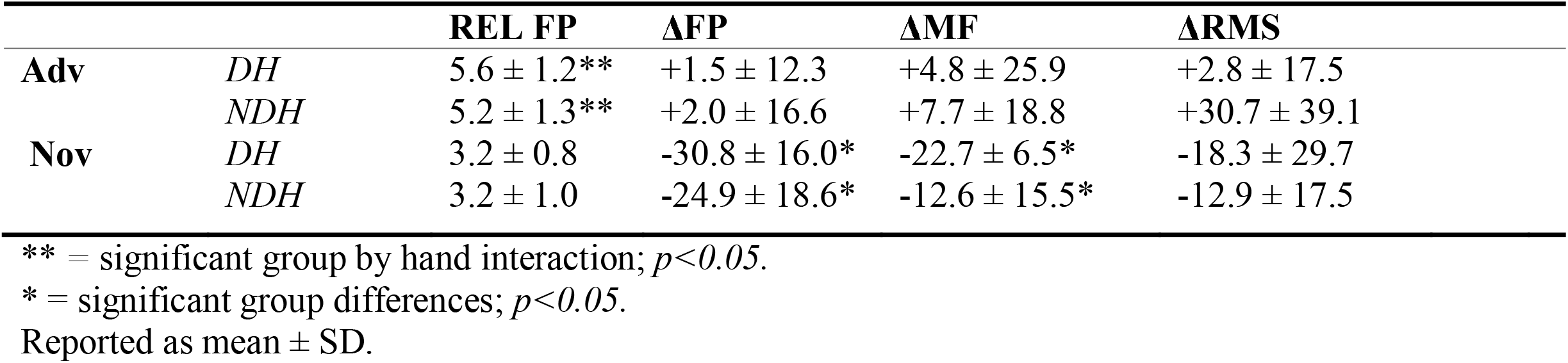
Change in finger force production and muscle activity (n=15)

|  |  | **REL FP** | **ΔFP** | **ΔMF** | **ΔRMS** |
| --- | --- | --- | --- | --- | --- |
| **Adv** | *DH* | 5.6 ± 1.2** | +1.5 ± 12.3 | +4.8 ± 25.9 | +2.8 ± 17.5 |
|  | *NDH* | 5.2 ± 1.3** | +2.0 ± 16.6 | +7.7 ± 18.8 | +30.7 ± 39.1 |
| **Nov** | *DH* | 3.2 ± 0.8 | -30.8 ± 16.0* | -22.7 ± 6.5* | -18.3 ± 29.7 |
|  | *NDH* | 3.2 ± 1.0 | -24.9 ± 18.6* | -12.6 ± 15.5* | -12.9 ± 17.5 |

\*\* = significant group by hand interaction; $p < 0.05$ .\* = significant group differences; $p < 0.05$ .
Reported as mean ± SD.

### 3.4 Change in Force Production

A significant group effect was observed for ΔFP (*F*_1_*= 16.1, p*=0.001). The advanced group experienced no reduction in FP, while the novice group’s FP significantly decreased from MVIC_PRE_ to MVIC_POST_ by 27.8 ± 16.7%.

### 3.5 Change in Muscle Activity

A significant main group effect was observed for FDS ΔMF (*F*_1_*= 15.1, p=0.002*), but not ΔRMS (*F*_1_*= 2.56, p=0.135*). The advanced group experienced no change from baseline in ΔMF, but the novice group’s ΔMF decreased significantly by 17.6 ± 10.9%. Neither advanced nor novice participants experienced a change in RMS. [Figure 2 near here].

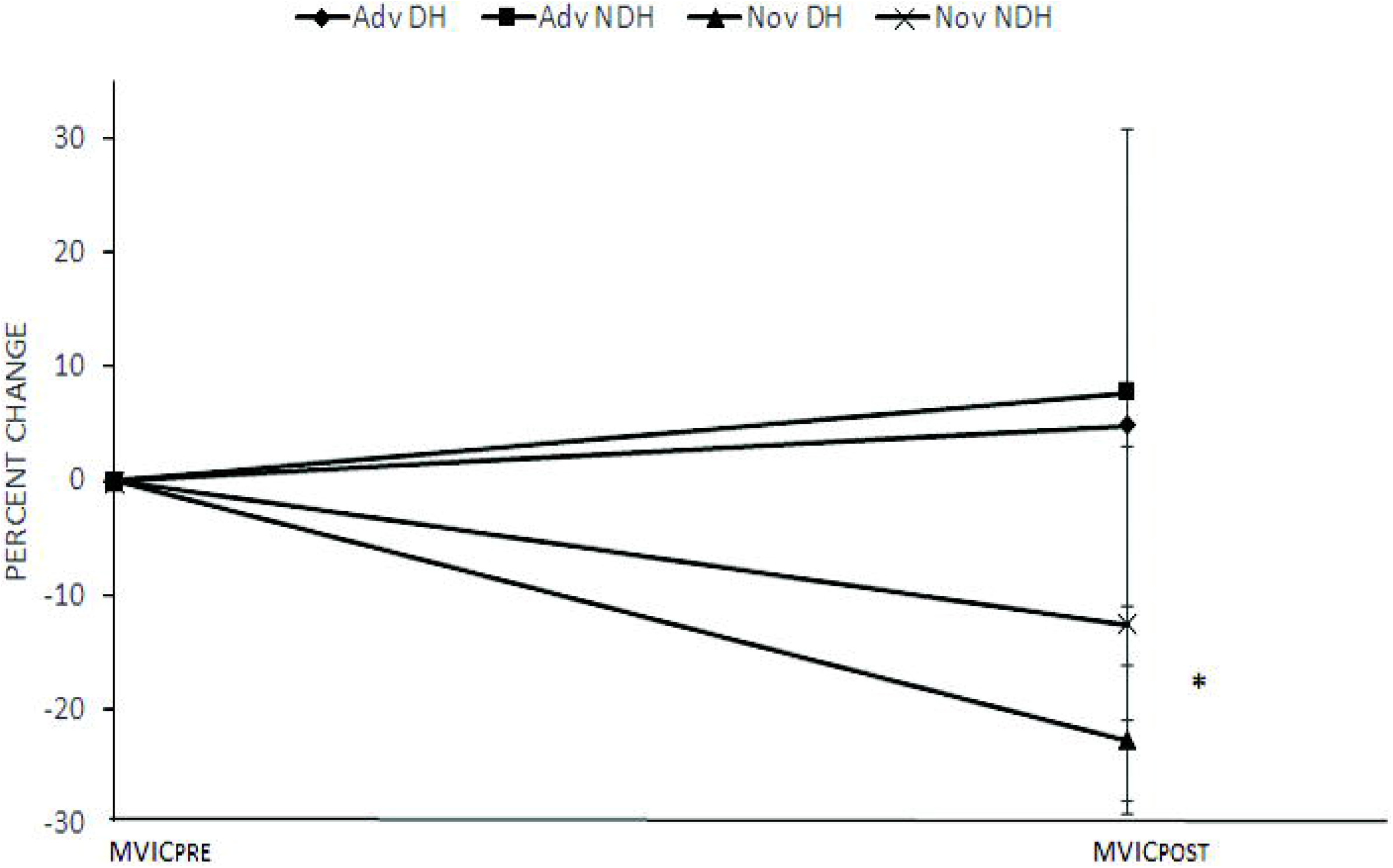

## 4. Discussion

The purpose of this study is to measure changes in finger force production and muscle activity in advance and novice sport rock climbers during a climbing protocol on a treadwall system. The results support our hypothesis that trained climbers will have a greater TTF, REL FP but lesser ΔFP and ΔMF over time when compared to the novices. While previous literature established differences between advanced climbers and novice/untrained counterparts during fatigue-inducing protocols, much of the research lacked sport-specificity. The current study utilizes a novel protocol, combining a mounted force plate to measure maximal finger force production with a treadwall system to fatigue climbers in a controlled, sport-specific setting.

### 4.1 Time to Fatigue

Previous research demonstrates trained climbers have greater localized endurance during both continuous and intermittent fingertip contractions against a force plate at fixed percentages of the subject’s MVIC.^9, 10, 15^ In the current study, advanced climbers also exhibited greater finger flexor endurance during actual bouts of climbing. This may be attributed to years of training, including metabolic adaptations and psychological conditioning of the forearm musculature. It has been suggested that forearm and whole body aerobic capacity can be used to predict climbing performance.^2, 24^ Specifically, advanced climbers have shown greater oxygenation capacity in the forearm musculature, leading to less reliance on anaerobic metabolism and a longer climb time. While muscle oxygenation capacity was not measured in the present study, a higher sport-specific aerobic capacity could account for differences in TTF between ability groups.

### 4.2 Force Production

Advanced climbers had greater finger force production relative to body weight (REL FP) during both pre- and post-climbing conditions compared to novice climbers (see Figure 2). Previous literature also described similar findings. Grant et. al, MacLeod et al., Phillippe et al., and Fryer et al. had comparable results, reporting trained climbers producing between 5.2 and 7.2 N/kg BW on a similarly mounted force plate.^2, 5, 13, 17^ In the current study, advanced climbers produced an average of 5.4 N/kg BW. Previous studies also reported a trend of increasing maximal FP with experience level, illustrating the importance of strength-to-weight ratio in high-performance free climbing. ^5-7, 9-11, 13^

When comparing ΔFP from MVIC_PRE_ to MVIC_POST_, the advanced group experienced no decline in force production after 30 minutes of climbing, whereas the novice group experienced significant decline. No direct comparisons to previous research can be made due to differences in protocol. However, Limonta et al. and Phillipe et al. reported similar findings during exhaustive simulated climbing protocols using sustained contractions against a force plate at 80% MVIC and intermittent contractions at 40% MVIC, respectively.^3, 17^ Both research groups found that trained climbers maintained higher levels of finger force production compared to novice/non-trained subjects. In the present study, the novice subjects’ FP tapered by 24-30% (and as much as 61%) at the point of failure during treadwall climbing. The advanced group, however, experienced no statistical decrease in FP after 30 min. of treadwall climbing. These findings again may suggest metabolic differences between advanced and novice climbers, possibly pointing to greater reliance on aerobic energy pathways in the forearm musculature.

### 4.3 EMG Data

Electromyographic data collected from the FDS supports the hypothesis for ΔMF, but not ΔRMS. No statistically significant ΔMF was observed in the advanced group, however the novice group’s MF decreased by 22% and 12% in the DH and NDH, respectively. (Fig. 3) Median frequency findings agree with Vigoroux & Quaine, who found elite-level climber’s MF decreases less quickly during fatiguing contractions when compared to non-trained counterparts.^11^ Previous research also demonstrates a correlation between MF and contraction intensity, relating to the recruitment of larger motor units with higher conduction velocities.^16^ It has been suggested that greater vasodilatory capacity of the forearm musculature could allow for greater recovery between contractions, thereby reducing the build-up of metabolites and “inducing a smaller change in the action potential velocity.”^12^ The lack of significant results in RMS could be attributed to the decline in overall fiber recruitment and reliance on Type I fibers for force production. More research should be devoted to activation patterns during fatigued states in rock climbers of all ability groups.

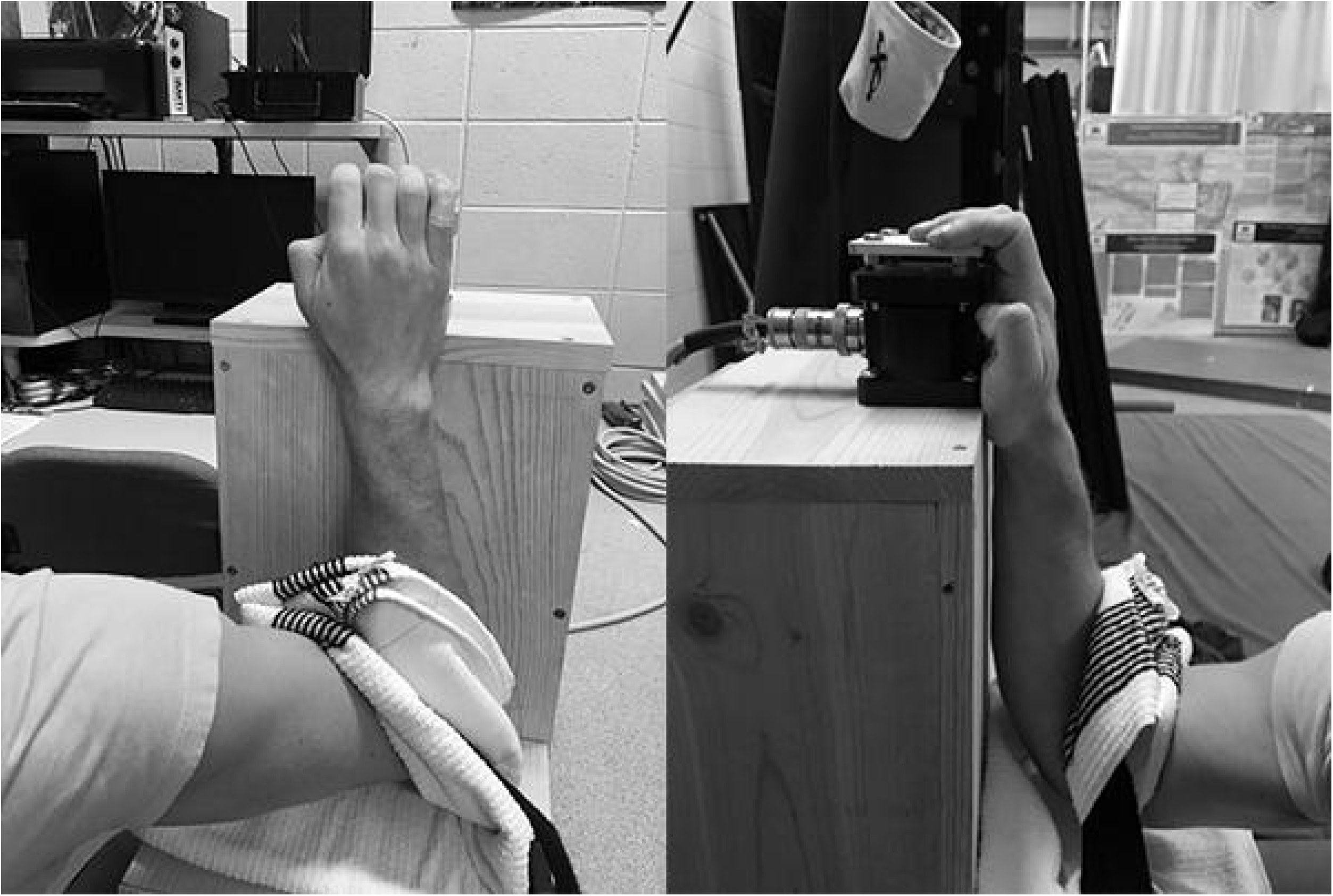

## 5. Conclusion

In this paper, we demonstrate differences between advanced and novice rock climbers using a treadwall system and mounted force plate. Rock climbing is a peculiar sport, requiring finger flexor force production, endurance and specific hand configurations not utilized in other grip strength-dependent sports like weightlifting. Previous climbing-related research used non-climbing means to induce fatigue in subject. Results of the present study build upon this literature, demonstrating advanced climbers have greater resistance to fatigue during actual bouts of climbing. Moreover, compared to the novice group, advanced climbers displayed greater finger force production relative to body weight and saw lesser decline in force production and MF. The results are significant, as it confirms previous findings and provides a new, sport-specific protocol for future rock climbing research.

Future studies may utilize a similar protocol to analyze the effect of supplementation, training intervention or further investigate climbing-specific metabolic capacities. Moreover, the force plate and treadwall system together could be used to identify differences between climbing subdisciplines.

## Data Availability

All data referred to in the manuscript is available to the public if requested.

## Acknowledgements

Thank you to Dr. Sophie Cowling, Dr. Tony Chang and Merrilee Thomas for their technical assistance and to the subjects for their participation in this study.

## Author Contributions

PF contributed in the design and implementation of the study protocol, data collection, data analysis and final manuscript writing. JB contributed in design of the protocol and editing of the final manuscript. JS contributed in the design and implementation of the study protocol, editing of the final manuscript and overall supervision.

## References

1. Roper S. Camp 4: Recollections of a Yosemite Rock Climber. The Mountaineers Books; 2013.

2. Fryer S, Stoner L, Lucero A, et al. Haemodynamic kinetics and intermittent finger flexor performance in rock climbers. International journal of sports medicine. 2015;36(02):137–142.

3. Philippe M, Wegst D, Müller T, Raschner C, Burtscher M. Climbing-specific finger flexor performance and forearm muscle oxygenation in elite male and female sport climbers. European journal of applied physiology. 2012;112(8):2839–2847.

4. Horst E. Training for Climbing: The Definitive Guide to Improving Your Performance. Rowman & Littlefield; 2016.

5. Grant S, Hynes V, Whittaker A, Aitchison T. Anthropometric, strength, endurance and flexibility characteristics of elite and recreational climbers. Journal of sports sciences. 1996;14(4):301–309.

6. Mermier CM, Janot JM, Parker DL, Swan JG. Physiological and anthropometric determinants of sport climbing performance. British journal of sports medicine. 2000;34(5):359–365.

7. Sheel A. Physiology of sport rock climbing. British journal of sports medicine. 2004;38(3):355–359.

8. Watts P, Newbury V, Sulentic J. Acute changes in handgrip strength, endurance, and blood lactate with sustained sport rock climbing. The Journal of sports medicine and physical fitness. 1996;36(4):255–260.

9. Vigouroux L, De Monsabert BG, Berton E. Estimation of hand and wrist muscle capacities in rock climbers. European journal of applied physiology. 2015;115(5):947–957.

10. Grant S, Shields C, Fitzpatrick V, et al. Climbing-specific finger endurance: a comparative study of intermediate rock climbers, rowers and aerobically trained individuals. Journal of Sports Sciences. 2003;21(8):621–630.

11. Vigouroux L, Quaine F. Fingertip force and electromyography of finger flexor muscles during a prolonged intermittent exercise in elite climbers and sedentary individuals. Journal of sports sciences. 2006;24(2):181–186.

12. Quaine F, Vigouroux L. Maximal resultant four fingertip force and fatigue of the extrinsic muscles of the hand in different sport climbing finger grips. International journal of sports medicine. 2004;25(08):634–637.

13. MacLeod D, Sutherland D, Buntin L, et al. Physiological determinants of climbing-specific finger endurance and sport rock climbing performance. Journal of sports sciences. 2007;25(12):1433–1443.

14. Watts PB, Drobish KM. Physiological responses to simulated rock climbing at different angles. Medicine & Science in Sports & Exercise. 1998;30(7):1118–1122.

15. Cram JR. Introduction to Surface Electromyography. Aspen Publishers; 1998.

16. Esposito F, Limonta E, Cè E, Gobbo M, Veicsteinas A, Orizio C. Electrical and mechanical response of finger flexor muscles during voluntary isometric contractions in elite rock-climbers. European journal of applied physiology. 2009;105(1):81..

17. Limonta E, Cè E, Gobbo M, Veicsteinas A, Orizio C, Esposito F. Motor unit activation strategy during a sustained isometric contraction of finger flexor muscles in elite climbers. Journal of sports sciences. 2016;34(2):133–142.

18. Watts PB, Jensen RL, Gannon E, Kobeinia R, Maynard J, Sansom J. Forearm EMG during rock climbing differs from EMG during handgrip dynamometry. International Journal of Exercise Science. 2008;1(1):2.

19. Schweizer A, Furrer M. Correlation of forearm strength and sport climbing performance. Isokinetics and Exercise Science. 2007;15(3):211–216.

20. Draper N, Dickson T, Blackwell G, et al. Self-reported ability assessment in rock climbing. Journal of sports sciences. 2011;29(8):851–858.

21. Draper N, Simon F, David W, et al. Reporting climbing grades and grouping categories for rock climbng. Isokinetics and exercise science. 2011;19:273-280. doi:10.3233/IES20110424

22. Blackwell JR, Kornatz KW, Heath EM. Effect of grip span on maximal grip force and fatigue of flexor digitorum superficialis. Applied ergonomics. 1999;30(5):401–405.

23. Watts PB, Jensen RL, Moss DM, Wagensomer JA. Finger strength does not decrease with rock climbing to the point of failure. Medicine & Science in Sports & Exercise. 2003;35(5):S264.

24. Fryer SM, Giles D, Palomino IG, de la O Puerta A, España-Romero V. Hemodynamic and cardiorespiratory predictors of sport rock climbing performance. The Journal of Strength & Conditioning Research. 2018;32(12):3534–3541.

